# An integrated deep mutational scanning approach provides clinical insights on *PTEN* genotype-phenotype relationships

**DOI:** 10.1101/2019.12.18.19015297

**Authors:** Taylor L. Mighell, Stetson Thacker, Eric Fombonne, Charis Eng, Brian J. O’Roak

**Author notes:** Correspondence; @CharisEngMDPhD. Correspondence; @TheRealDrOLab. These authors contributed equally to this work (Taylor L. Mighell, Stetson Thacker).

## Abstract

Germline variation in *PTEN* results in variable clinical presentations, including benign and malignant neoplasia and neurodevelopmental disorders. Despite decades of research, it remains unclear how *PTEN* genotype is related to clinical outcomes. In this study, we combined two recent deep mutational scanning (DMS) datasets probing the effects of single-amino acid variation on enzyme activity and steady-state cellular abundance with the largest well-curated clinical cohort of *PTEN*-variant carriers. We sought to connect variant-specific molecular phenotypes to the clinical outcomes of individuals with *PTEN* variants. We found that DMS data partially explain quantitative clinical traits, including head circumference and Cleveland Clinic (CC) score, which is a semi-quantitative surrogate of disease burden. We built a logistic regression model using DMS and CADD scores to separate clinical *PTEN* variation from gnomAD control-only variation with high accuracy (AUC = 0.892). Using a survival-like analysis, we identified molecular phenotype groups with differential risk of early-onset as well as lifetime risk of cancer. Finally, we identified classes of DMS-defined variants with significantly different risk levels for classical hamartoma-related features (odds ratios range of 4.1-102.9). In stark contrast, the risk for developing autism or developmental delay does not significantly change across variant classes (odds ratios range of 5.4-12.4). Together, these findings highlight the potential impact of combining DMS datasets with rich clinical data, and provide new insights that may guide personalized clinical decisions for *PTEN*-variant carriers.

## Introduction

Germline mutation of the tumor suppressor gene phosphatase and tensin homolog (*PTEN* [MIM: 601728]) manifests with variable and complex phenotypes, including macrocephaly (with increased occipital-frontal circumferences [OFC]), benign hamartomas affecting all three germ layers, malignant neoplasia across multiple tissues, and neurodevelopmental abnormalities, including autism spectrum disorder (ASD).^1,2^ This heterogeneity is reflected clinically with germline *PTEN* mutations found in variable subsets of defined syndromes, including Cowden syndrome and Bannayan-Riley-Ruvalcaba syndrome (CWS1 and BRRS [MIM: 158350]), as well as macrocephalic ASD (MAS [MIM: 605309]), among others.^2–5^ Collectively, these syndromes have been termed PTEN hamartoma tumor syndrome (PHTS) when a germline *PTEN* variant is identified.^1,2^

The dramatic variability of these clinical presentations has sparked efforts to correlate *PTEN* variants with clinically-relevant phenotypic classes. However, *PTEN* variants resist simple classification approaches based on secondary domain clustering or variant type. Recently, the mapping of a limited subset of germline *PTEN* variants onto the three-dimensional, crystal structure failed to reveal a distinct pattern of distribution between ASD-or cancer-predisposition-associated variants.^6^ Classification efforts have also been impacted by limited sample sizes in terms of both functional data and *PTEN* variant cohorts.^7,8^ Additionally, the PTEN protein has multiple functional roles in the cell apart from lipid phosphatase activity, which may also play a role in this phenotypic complexity.^2,9–11^

These challenges have prompted recent creative high-throughput methods to functionally measure the molecular phenotypes for thousands of nonsynonymous *PTEN* variants, collectively termed deep mutational scanning (DMS).^8,12^ We previously reported the effect of nearly all *PTEN* nonsynonymous variants on lipid phosphatase activity by utilizing a humanized yeast assay where lipid phosphatase activity was linked to cell survival (so called fitness score).^8^ These data demonstrated that the solvent exposure of a wild-type residue is a critical determinant of mutational tolerance for lipid phosphatase fitness, with solvent exposed residues being much more tolerant to mutation. As expected, PTEN lipid phosphatase activity was generally intolerant to mutation in the catalytic pocket and phosphatase domain, though not without exception. Further, in line with suggestions from prior more limited functional studies^7^, *PTEN* missense variants associated with ASD tended to retain partial lipid phosphatase activity.^8^

In a second independent study, the effect of ∼54% of all *PTEN* nonsynonymous variants on the steady-state cellular protein abundance were estimated using fluorescently tagged PTEN variant proteins (so called abundance score). It was observed that PTEN abundance is, in part, explained by thermodynamic stability and cell-membrane interactions of a given variant. While variant abundance inversely correlates with pathogenicity, notable exceptions are putative dominant-negative PTEN variants, which are highly stable but catalytically inactive^12^.

While these two DMS studies have added essential insights into the effect of *PTEN* variants on protein function, they were limited in their clinical analyses as both relied on previously published clinical reports and ClinVar database^13^ variants with varying degrees of validation and phenotypic description. In this study, to further uncover *PTEN* genotype-phenotype relationships and clarify patient risk for these diverse clinical presentations, we integrated these datasets with the largest, prospectively accrued and comprehensively clinically characterized cohort of *PTEN* variant-positive individuals (Cleveland Clinic [CC] cohort). These analyses demonstrate that molecular phenotypes associate with quantitative clinical traits, can be combined to improve pathogenicity prediction, delineate differential lifetime cancer risk, and indicate unexpected risk ratio relationships for neurodevelopmental and hamartoma-associated phenotypes.

## Materials and Methods

### *PTEN* Variant Function Data and Imputation

We made use of two DMS datasets in this study.^8,12^ Briefly, fitness scores were previously determined by assessing a *PTEN* variant’s ability to reverse toxicity by means of phosphatidylinositol (3,4,5)-triphosphate (PIP_3_) dephosphorylation in a humanized yeast system^14^ that expresses a hyperactive kinase.^8^ High confidence fitness scores were previously generated for 86% of all variants and a random forest algorithm was used to impute fitness scores for the remaining unmeasured variants.^8^ Abundance scores were previously determined by measuring the steady-state level of *PTEN* variants using the VAMP-Seq assay in human cells.^12^ Abundance scores were generated for 54% of all variants. Using a random forest framework similar to what was used to impute fitness scores, here, we imputed abundance scores for the remaining unmeasured variants (Figure S1). Modeling was implemented in Scikit-learn version 0.19.0 (sklearn.ensemble.RandomForestRegressor, n_estimators=500, criterion= “mse”, max_features=0.33, random_state=0, oob_score=True). We trained a model on position average, i.e. the average score of all other substitution variants at that amino acid position and the n-1 and n+1 positions. If there were no measured variants at the n-1, n, or n+1 positions, we included the n-2 and n+2 positions. We then iteratively trained models incorporating features in the order of their importance until the model Pearson correlations plateaued. Feature importance was calculated as the relative increase in error upon random permutation of a feature. The final model was assessed using 10-fold cross validation, i.e. train the model on 90% of data and test on the remaining 10% (Figure S1 and Table S1). Finally, we used the final model trained on all measured abundance scores to predict all unmeasured variants.

We classified the full set of missense protein variants (measured and imputed) as wildtype-like, hypomorphic, or truncation-like for fitness and abundance scores (Figure S2, Table S1). For fitness score, we considered variants wildtype-like if they were within the 2.5 and 97.5 percentile of synonymous wildtype fitness scores (Figure S2C-D). We considered variants truncation-like if their fitness scores were within the 2.5 and 97.5 percentile of nonsense variants at positions 1-350, excluding the regulatory tail because nonsense mutations in the tail are not damaging. We considered variants hypomorphic for fitness score if they were between the wildtype-like and truncation-like bounds.

We classified wildtype-like variants similarly for abundance score with a slight adjustment to the distribution boundaries (Figure S2C-D). Because the abundance score distribution tails were larger than the fitness score distribution tails, we defined the bounds as the 5 and 95 percentile of synonymous wildtype distribution. We considered variants to be truncation-like for abundance score if they were within the 5 and 95 percentile of nonsense variants at positions 30-300, in order to exclude artifacts of variants near the protein termini.

Wildtype *PTEN* nucleotide and corresponding amino acid sequence were obtained from GenBank (NM_000314.6) and GenPept (NP_000305). PTEN partial crystal structure was obtained from PDB (1D5R).

### *PTEN* Population Variants from GnomAD

Data from the controls-only subset was downloaded from gnomAD v2.1^15^ on January 10, 2019 (Table S2). For the cancer incidence analysis and the clinical outcomes odds ratio analyses, we included all controls-only gnomAD nonsynonymous variants (e.g., missense, nonsense, and indel frameshift). For the pathogenic vs. benign analysis, we considered only gnomAD missense variants with the exceptions of p.Arg173His and p.Lys289Glu, which are classified as pathogenic or likely pathogenic in ClinVar; and p.Asp268Glu, which occurs at a frequency greater than an order of magnitude over most other variants.

### Cleveland Clinic *PTEN* Cohort

This study was performed in accordance with the IRB# 8458 protocol “Molecular Mechanisms Involved in Cancer Predisposition” substudy PTEN, which has been approved by the Cleveland Clinic Institutional Review Board for Human Subjects’ Protection, and conducted with informed consent and in accordance with the World Medical Association Declaration of Helsinki. The CC cohort consists of 256 prospectively accrued individuals with germline *PTEN* nonsynonymous variants (145 missense and 111 nonsense variants, Table 1 and Tables S3-S4). Genotype information concerning each patient’s germline *PTEN* variant, demographic, and clinical data were also included. Collection and validation of clinical phenotypes were performed by experienced clinical personnel as detailed in a previous study.^16^ Demographic information includes the age at last follow-up, sex, and age at diagnosis for various clinical phenotypes, including: macrocephaly, neurodevelopmental pathologies including ASD and developmental delay (DD), and several different types of benign and malignant neoplasia. Adult individuals in the cohort were given a CC score, which is a semi-quantitative estimation of the probability of having a germline *PTEN* mutation based on clinical phenotypes. For example, a score of 15 indicates a 10% probability. The CC score also serves as a surrogate measure of burden of disease with larger scores indicating increasing disease burden and/or younger ages of onset. However, the scoring is only applicable/validated for the adult population (includes individuals 18 and older).^16^OFC z-scores were calculated using published, age-indexed tables.^17^

**Table 1.** CC cohort of individuals with germline nonsynonymous variation in *PTEN*.

| Phenotype | Missense |  | True Truncations |  | Total |  |  |
| --- | --- | --- | --- | --- | --- | --- | --- |
|  | All N (%) | % Male | All N (%) | % Male | All N (%) | % Male | % Mis. |
| All | 145 (100) | 40.7 | 111<sup>a</sup> (100) | 49.5 | 256 (100) | 44.5 | 56.6 |
| ASD/DD | 23 (15.9) | 78.3 | 6 (5.41) | 100.0 | 29 (11.3) | 82.8 | 79.3 |
| ASD/DD & PHTS | 32 (22.1) | 68.8 | 24 (21.6) | 75.0 | 56 (21.9) | 71.4 | 57.1 |
| PHTS | 90 (62.1) | 21.1 | 80 (72.1) | 38.8 | 170 (66.4) | 29.4 | 52.9 |

<sup>a</sup> One individual did not have qualifying symptoms for any phenotype group.

### Logistic Regression Modeling for Pathogenic *PTEN* Variation

To test the accuracy of models with all combinations of features (e.g., fitness scores, abundance scores, and CADD scores), the optimal regularization parameters (L1 vs. L2 regularization and regularization strength) for each feature combination were determined using the GridSearchCV function within scikit learn. The best performing model as judged by area under the curve analysis used fitness scores, abundance scores, and CADD scores, with a L1 regularization strength of 1/2.78.

### Cancer Incidence and Survival Analysis

For fitness and abundance score analyses, we classified all individuals from the CC cohort and gnomAD into wildtype-like missense, hypomorphic missense, truncation-like missense, or true-truncation (i.e. nonsense or frameshifting) groups (Figure S2). For the combined molecular score analysis, we used the hypomorphic cutoff to designate variants as fitness or abundance plus or minus (i.e., -1.11 for fitness score, 0.71 for abundance score). We assumed the gnomAD individuals were cancer free. The observation period for each subject was set from birth to age at last clinical follow-up/information. For 26 of the 164 gnomAD individuals, we could unambiguously determine their age range by linking to data provided in the full gnomAD v2.1 variant call file. Since these data were provided in five-year increments, we randomly selected a single year from this range for each individual. For the rest of the gnomAD control cohort, we imputed age by using the whole dataset age distribution that fits approximately a normal distribution with mean of 55 and standard deviation of 15. For each individual, we assigned the mean of five ages (rounded to the nearest integer) randomly generated from this distribution. We rounded to the nearest integer for consistency with the rest of the age data. Differences in cancer incidence between the genotype groups were compared using the Kaplan-Meyer method and log-rank test. Analyses were performed for overall cancer incidence and individuals were right-censored at age at cancer or age at last follow-up. Significant group differences were then examined using pair-wise comparisons. In order to detect potential differences in early onset cancer incidence, survival curves were further compared at age 35 with right-censoring occurring at age of early onset (<35) cancer or age 35 otherwise.

### Calculating Odds Ratios for Clinical Outcomes

Although all individuals with an identified germline, pathogenic *PTEN* variant are clinically classified as belonging to the overarching classification of PHTS, we have developed clinical subgroupings with differing presentations in order to enable genotype-phenotype analyses in this study. An individual was considered ASD/DD positive if they presented with ASD, DD, variable delay, or intellectual disability. An individual was considered PHTS positive if they presented hamartomatous features including any of the following: benign or malignant tumors, mucocutaneous lesions, arteriovenous malformation, lipomas, goiter, or uncommon skin lesions. Individuals with the common skin findings of skin tags, café-au-lait marks, or penile freckling in isolation, meaning without another hamartomatous feature, were not included in the PHTS group. Individuals who displayed both the neurodevelopmental and hamartomatous features were placed in the ASD/DD & PHTS grouping. Individuals with frameshifting and nonsense mutations were treated as true truncations. We used IBM SPSS statistical software to perform logistic regression modeling on ASD/DD or PHTS outcomes, using molecular phenotypes as exposures.

## Results

### Distribution of Missense Variation across the Primary and Crystal Structures of PTEN

In order to examine *PTEN* genotype-phenotype relationships, we prospectively accrued a cohort of individuals with germline nonsynonymous variation in *PTEN* (Table 1). Previously, PHTS has been used as an umbrella term specifically for classically defined *PTEN*-related disorders (e.g., Cowden syndrome and Bannayan-Riley-Ruvalcaba syndrome).^18^ Subsequently, as the phenotypic spectrum of *PTEN* mutations expanded, PHTS became a descriptor for all clinical presentations associated with germline *PTEN* variation.^2^ In order to explore potential differences between ASD/DD related phenotypes and those associated with hamartoma/cancer phenotypes, we operationally grouped individuals with the classic hamartoma-related *PTEN* features as PHTS, while individuals with largely neurodevelopmental clinical features were designated ASD/DD (Materials & Methods). Individuals with a combination of neurodevelopmental and hamartoma-related features were designated as a third group, ASD/DD & PHTS.

The cohort recapitulates the previously observed relative enrichment of ASD/DD phenotypes among those with missense as opposed to nonsense variation (16% vs 5%, Table 1).^11,19^ As a comparison, we also collated mutation data from control-only individuals in gnomAD, a database that aggregates sequencing studies.^15^ These individuals were assumed to be unaffected by *PTEN*-related disorders. We categorized missense variants by associated clinical group and then mapped variants to the primary/functional domain and crystal structures of PTEN, including the variants catalogued in gnomAD (Figure 1A-B). The clinical missense variants cluster most heavily in the dual-specificity phosphatase domain (residues 1-178), and are depleted in the C2 domain & tail (residues 179-403), reflecting the importance of this protein domain to *PTEN* function (Figure 1A). Comparing all clinical variants with gnomAD variants demonstrates an enrichment of gnomAD variants in the C2 domain & tail, as compared to the phosphatase domain (p=7.9x10^−9^, Fisher’s exact test, Figure 1B). In contrast, and consistent with similar studies,^6^ the distributions of variants were similar across clinical outcomes (ASD/DD, PHTS, or ASD/DD & PHTS, Figure 1A). In 3D space, gnomAD variants are significantly more solvent exposed (i.e., exposed to the surface of the protein) than the group of all clinical variants (medians of 87.4% vs 7.8%, p=1.28x10^−18^; Mann-Whitney U-test, Figure 1B). Variation at solvent exposed positions is generally more tolerated because these variants are less likely to disrupt protein structure.^20^

**Figure 1.**
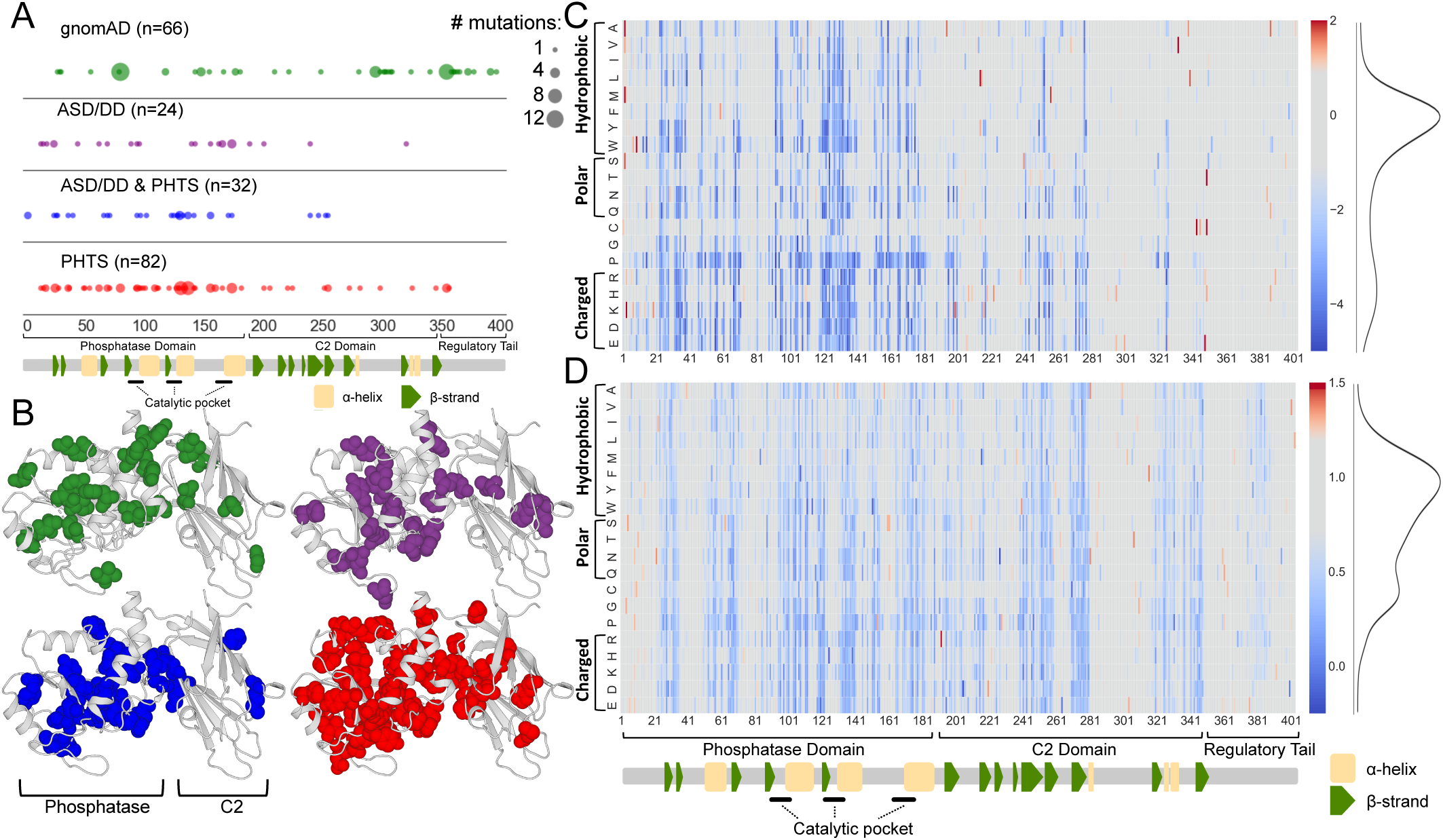
Overview of the datasets used in this study. (A) Diagram of PTEN primary protein structure with locations of PTEN missense variants found in the controls-only gnomAD population or associated with various clinical presentations in the CC cohort. The major protein domains and secondary structure assignments are indicated. Size of circle indicates number of different amino acid variants at that position. (B) Diagram of PTEN 3D crystal structure with locations of PTEN missense variants found in the controls-only gnomAD population or associated with various clinical presentations in the CC cohort. The C-terminal tail was not solved in the crystal structure and therefore variants falling in this region are not shown. Color of spheres indicates same groups as shown in (A). (c) Lipid phosphatase fitness scores are displayed as a heatmap, with blue corresponding to damaging variants (i.e. low lipid phosphatase fitness or low abundance), gray corresponding to wildtype-like, and red corresponding to putative fitness or abundance increasing variants (Materials & Methods). The distributions of all missense variants are shown as smoothed histograms on the right. (D) Cellular abundance scores displayed as in (C).

### Visualization and Imputation of Molecular Phenotypes of *PTEN*

We hypothesized that variant level molecular phenotype data might uncover new genotype-phenotype associations, which are more complex than simple clustering of clinical outcomes in primary or tertiary protein sequence space. We aggregated molecular phenotype information derived from recent DMS studies on the effect of thousands of variants on PTEN protein function, including inferred lipid phosphatase activity (i.e., fitness score) and steady-state protein stability (i.e., abundance score).^8,12^ Previously, we demonstrated that by using a random forest-based machine learning modeling approach, fitness scores of variants withheld from model training could be imputed with high accuracy. The model incorporated the position average effect of variants missing from a nearly complete DMS dataset (86% saturation) with biophysical, biochemical, and evolutionary data. Therefore, using the imputations from this model, we previously constructed a comprehensive lipid phosphatase functional map of fitness scores (Figure 1C).

We reasoned that a similar strategy might be used, again with high accuracy, even for less complete DMS datasets. We developed a similar modeling strategy for the protein abundance DMS dataset, which was at ∼54% saturation. Cross validation showed the best performing model could accurately predict withheld abundance scores with an accuracy similar to biologic replicates (Pearson r = 0.75, Figure S1). Therefore, using this approach, we imputed abundance scores for all missing missense variants (Figure 1D and Table S1). Combined, these complete datasets represent estimates of the effect of any given *PTEN* missense variant on the lipid phosphatase activity and steady-state abundance of PTEN protein. All analyses presented here used the combination of high confidence measured and imputed scores.

Fitness scores are modestly correlated with abundance scores (Pearson’s r = 0.43, Figure S2E), suggesting some information overlap but that each assay is also capturing unique variant effects on protein function. We used the distribution of programmed truncating (nonsense) and synonymous variants in these assays to define truncation-like, hypomorphic, and wildtype-like missense variant categories (Materials and Methods, Figure S2A-D). The missense variants in both datasets are bimodally distributed, with the majority of variants having wildtype-like scores (Figures 1C-D and S2). We found that for both measures, variation in the phosphatase domain was generally more damaging, i.e. truncation-like or hypomorphic, than variation in the C2 domain or regulatory tail (fitness: 42% vs. 12%, abundance: 50% vs. 38%, Figure 1C-D).

### Fitness and Abundance Scores Explain Quantitative Clinical Traits

In an effort to link genotype to quantitative clinical phenotypes, we evaluated whether fitness and abundance scores of individuals’ *PTEN* missense variants could explain the degree of macrocephaly or phenotype burden as assessed by CC score for individuals over 18 (Materials and Methods). Molecular phenotype scores were evaluated numerically as well as using the defined functional categories (e.g., wildtype-like, hypomorphic, and truncation-like). We found a logarithmic relationship between fitness score and head size measured by OFC, with z-scores plateauing around the hypomorphic cutoff (Figure 2A, left). Accordingly, we found a significant difference in OFC between the population of wildtype-like variants and truncation-like as well as hypomorphic variants (p = 4.3x10^−5^ and 6x10^−4^, respectively, Mann-Whitney U-test), but no difference between hypomorphic and truncation-like variant fitness scores (Figure 2B, left). We also observed a logarithmic relationship between OFC and abundance score (Figure 2A, right). Treating abundance as a categorical variable revealed significant differences between wildtype-like and both truncation-like and hypomorphic variants (p = 0.02 and 0.01, respectively, Mann-Whitney U-test). Similar to the fitness score, there was no difference between the distribution of truncation-like and hypomorphic variants (Figure 2B, right).

**Figure 2.**
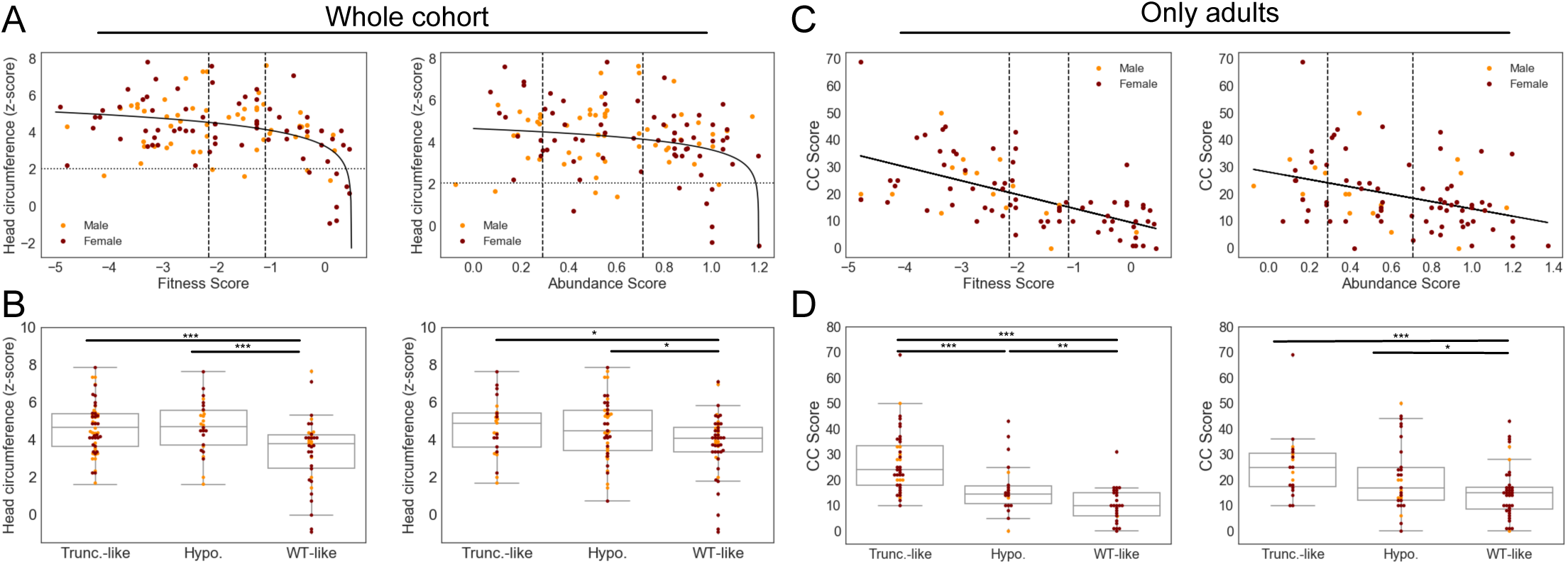
Relationships between molecular phenotype scores and quantitative clinical traits for missense variants. (A) Occipital frontal circumference (OFC, z-score) plotted as a function of continuous fitness score or abundance score for all individuals with missense variants. Males are shown as orange, females are shown as maroon. Vertical dashed lines indicate the hypomorphic and truncation-like cutoffs at -1.11 and -2.15, respectively. Horizontal dashed line indicates threshold for macrocephaly (z-score = 2.054). Solid lines indicate logarithmic curves fit to the data. (B) Box plot of OFC z-scores for all individuals with missense variants with fitness or abundance scores in the wildtype-like, hypomorphic, or truncation-like ranges. (C) score for adults with missense variants as a function of continuous fitness or abundance scores. Analyses are restricted to adults as CC score is not valid for individuals under 18. Vertical dashed lines indicate the hypomorphic and truncation-like cutoffs at - 1.11 and -2.15, respectively. Solid lines indicate linear curves fit to the data. (D)Box plot of CC scores for adults with missense variants with fitness or abundance scores in the wildtype-like, hypomorphic, or truncation-like ranges. *p<0.05; **p<0.01; ***p<0.001.

In our analysis of phenotype burden, we found a significant linear relationship between missense variant fitness score and CC score (p = 3.7x10^−10^), with fitness score explaining 37% of the variation in CC score (Figure 2C, left). Similarly, treating fitness score as a categorical variable, we found that more damaging groups of variants had distributions shifted toward higher (more severe) CC scores (Figure 2D, left). CC scores for truncation-like variants were significantly higher than those of hypomorphic variants (p = 2.5x10^−4^). Additionally, CC scores for hypomorphic variants were in turn significantly higher than those of wildtype-like variants (p = 9.2x10^−3^).

Alternatively, for abundance scores, while a significant linear relationship exists between CC score and abundance score (p = 3.2x10^−4^), it explains only 14% of the variation in CC score (Figure 2G). Likewise, when treating abundance score as a categorical variable, more modest trends were observed compared to the trends for fitness score. The abundance scores for truncation-like variants trend toward higher CC scores than hypomorphic variants (p = 0.08), while hypomorphic variants are nominally higher than wildtype-like (p = 0.045) variants. Truncation-like variants are significantly different from wildtype-like variants (p = 6.7x10^−4^; Figure 2D). Combined, these results underscore the potential for molecular phenotypes to partially explain clinical outcomes.

### Molecular Phenotype Data Accurately Distinguishes Likely Pathogenic from Benign Variation

We next examined whether molecular phenotype data could be utilized to identify pathogenic or likely pathogenic missense variation in *PTEN*. Thus, we contrasted the CC cohort of known pathogenic *PTEN* variants to putatively benign, population *PTEN* variants catalogued in the gnomAD control-only individuals (Materials and Methods, Figure 3, Table S5). We found that variants from the CC cohort were predicted to be significantly more damaging by both fitness score (p = 6.5x10^−13^, Mann-Whitney U-test) and abundance score (p = 7.6x10^−6^, Figure 3A-B). As a comparison, CADD scores^21,22^ are also more damaging for the CC cohort group (p = 6.5x10^−8^, Figure 3C). Of these three predictors, fitness scores demonstrate the highest area under the receiver operating characteristic curve (AUC = 0.888, Figure 3D). While these predictors are correlated, the relationships are modest (Spearman rho = 0.52-0.59, Figure S3A), suggesting that multivariate models could yield improved performance. Therefore, we constructed logistic regression models using various combinations of the molecular phenotype data and CADD scores to find the model that most accurately discriminates the two groups (Figure S3B). We found the most accurate model utilized fitness, abundance, and CADD scores to predict likely pathogenicity variants with AUC = 0.892 (Figure 3D). These findings highlight the improvement in computational models of pathogenicity that can be gained from empirical molecular phenotype data.

**Figure 3.**
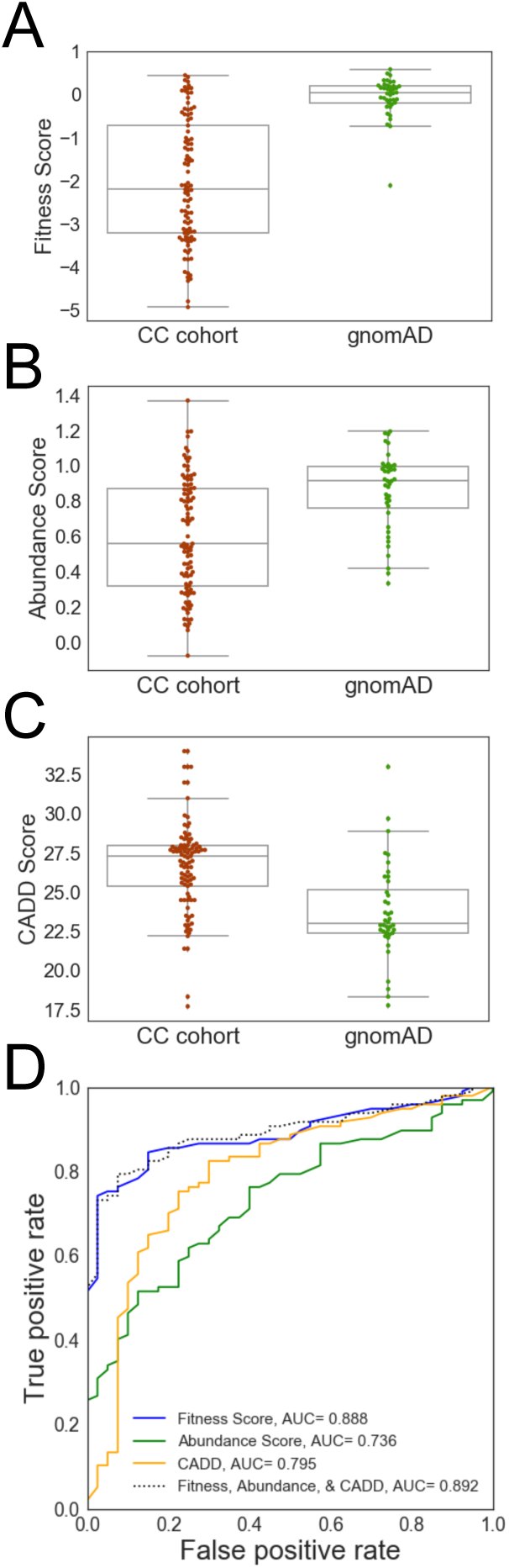
Molecular phenotypes discriminate clinical from gnomAD missense PTEN variation. These analyses are done at the variant level to prevent recurrent variants from biasing the results. Two known pathogenic variants were removed from the gnomAD list (Materials & Methods). (A) Box plots comparing fitness scores between variants found in the CC cohort versus gnomAD. (B) Box plots comparing abundance scores between variants found in the CC cohort versus gnomAD. (C) Box plots comparing CADD scores between variants found in the CC cohort versus gnomAD. (D) Receiver operator characteristic curves for individual predictors, as well as the optimized multivariate logistic regression using fitness, abundance, and CADD scores.

### Molecular Phenotypes Identify Subgroups with Distinct Cancer Susceptibility

While it is known that *PTEN* mutations dramatically increase the lifetime risk of developing specific cancers, we sought to understand whether molecular phenotypes could highlight functional classes of missense variants with differences in cancer susceptibility (Materials and Methods). As a comparison group, we included individuals with variants that are predicted to be truly truncating (e.g., nonsense and frameshifting). Survival functions were first compared between all classes of missense fitness or abundance scores and the true truncations, with pairwise comparisons of survival functions when significant differences were found (Figure 4A-B). In this analysis, cancer free status was considered as the survival criterion.

**Figure 4.**
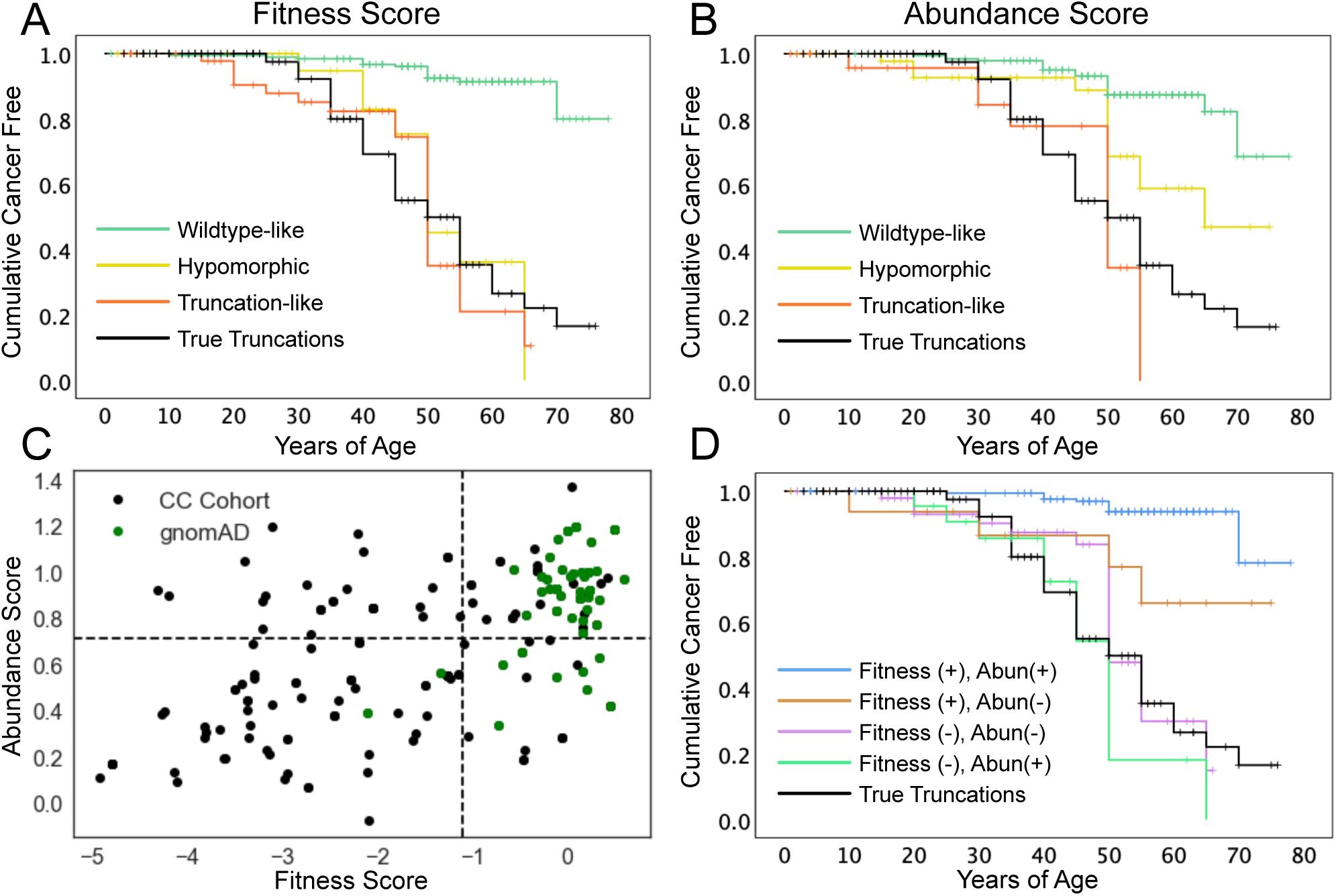
Effects of molecular phenotype on cumulative cancer incidence. (A) Survival-like analysis of individuals with different fitness score classes of *PTEN* missense or true-truncation (e.g. nonsense, frameshifting) variants. “Survival” here is defined as being cancer-free. Ticks represent right-censored individuals, i.e. the age at last follow-up. n=204 wildtype-like; 35 hypomorphic; 68 truncation-like, 114 true truncations. (B) Survival-like analysis of individuals with different abundance score classes of *PTEN* missense or true truncation (e.g. nonsense, frameshifting) variants. “Survival” here is defined as being cancer-free. Ticks represent right-censored individuals, i.e. the age at last follow-up. n=210 wildtype-like; 65 hypomorphic; 32 truncation-like, 114 true truncations. (C) All CC cohort and gnomAD individuals plotted as a function of fitness and abundance scores. Dotted lines indicate wildtype-like thresholds (-1.11 and 0.71 for fitness and abundance scores, respectively). (D) Survival-like analysis for individuals with *PTEN* missense variants falling in fitness/abundance quadrants or true truncating variants. “Survival” here is defined as being cancer-free. Ticks represent right-censored individuals, i.e. the age at last follow-up. n=29 fitness(-), abundance(+); 74 fitness(-), abundance(-); 23 fitness(+), abundance(-); 181 fitness(+), abundance(+), and 114 true truncations.

For fitness scores, survival functions were significantly different (p = 4.8x10^−24^, Log rank, Figure 4A and Table S6). Pairwise comparisons showed that all of the reduced fitness score categories survival functions were similar to each other and significantly different from the wildtype-like survival function (Table S6). Based on the shape of the survival functions, we hypothesized that there may be a difference in early on-set risk. Therefore, we conducted a subanalysis with right-censoring at age 35, which again showed significant overall differences (p = 3.0x10^−6^, Log rank). Pair-wise comparisons showed that these differences were driven by truncation-like and true truncations categories, each significantly deviated from wildtype-like variants (p = 9.0x10^−6^ and 2.8x10^−7^). The hypomorphic survival function appears to be intermediate between the groups. However, across this age range the hypomorphic function did not significantly differ from the wildtype-like function (p = 0.342), unlike the truncation-like and true truncation functions. Further supporting a potential intermediate early risk, when compared to both truncation groups the survival function for hypomorphic variants was suggestive of a differential cancer risk although the trend did not reach significance (p = 0.155 and 0.136, Figure 4A).

Variant classes defined by abundance scores also had significantly different survival functions (p = 2.1x10^−18^, Log rank, Figure 4B and Table S6). In contrast to the fitness scores, pairwise comparisons revealed a step-wise relationship for abundance scores, with hypomorphic missense variants conferring greater lifetime hazard than wildtype-like (p = 0.001), and both truncation-like missense and true truncations conferring greater hazard than the hypomorphic abundance class (p = 2.0x10^−11^ and 6.0x10^−14^, Figure 4B). Right-censoring at age 35 identified significantly different survival functions for the abundance-defined variant classes as well (p = 8.0x10^−6^). Similar to the fitness score analysis, these differences were driven by truncation-like and true truncations functions which were significantly different from wildtype-like between birth and age of 35 (p = 2.1x10^−5^ and 7.5x10^−7^, respectively). Again, these data were suggestive of the hypomorphic survival function being intermediate between wildtype-like (p = 0.06) and the truncation groups (p = 0.152 and 0.193), but none of the comparisons were significantly different.

We next leveraged the two-dimensional molecular phenotype data to separate missense variants into four categories based on deficiencies in fitness score, abundance score, or both. For this analysis, hypomorphic and truncation-like scoring variants were combined as the negative group for PTEN function for each score in order to keep adequate group sizes. Compared to CC Cohort, gnomAD individuals are enriched in the fitness positive, abundance positive quadrant (Figure 4C). The survival functions for these combined molecular phenotype defined groups were significantly different (p = 2.6x10^−24^, Log rank). Pairwise comparisons, showed variants retaining wildtype-like fitness (+) and abundance (+) have the lowest overall hazard and have a survival function that is significantly different from all other groups (Table S6). Fitness positive but abundant negative variants showed intermediate risk that was significantly different from both the wildtype -like and true truncations (p = 0.004 and 0.048, respectively). The remaining two classes are both fitness score deficient (abundance + or -) and are not significantly different from each other or the true truncations. These data suggest that combining multiple molecular phenotypes provides additional insights into cancer risk.

### Molecular Phenotypes Identify Distinct Risk Profiles for ASD/DD and PHTS Subgroupings

Understanding the molecular differences between the variants that associate with ASD/DD versus PHTS (especially cancer occurrence) outcomes is a critical goal for understanding PTEN pathobiology, which ultimately guides clinical management. Consistent with our and others’ previous findings,^7,8^ fitness scores of individuals in the ASD/DD group are less damaging than the PHTS positive groups (p = 5.5x10^−3^, 0.011, Mann-Whitney U, for ASD/DD vs ASD/DD & PHTS and ASD/DD vs PHTS, respectively, Figure 5A). However, there is no difference in abundance or CADD scores between the clinical phenotype groups (Figure 5B and data not shown).

**Figure 5.**
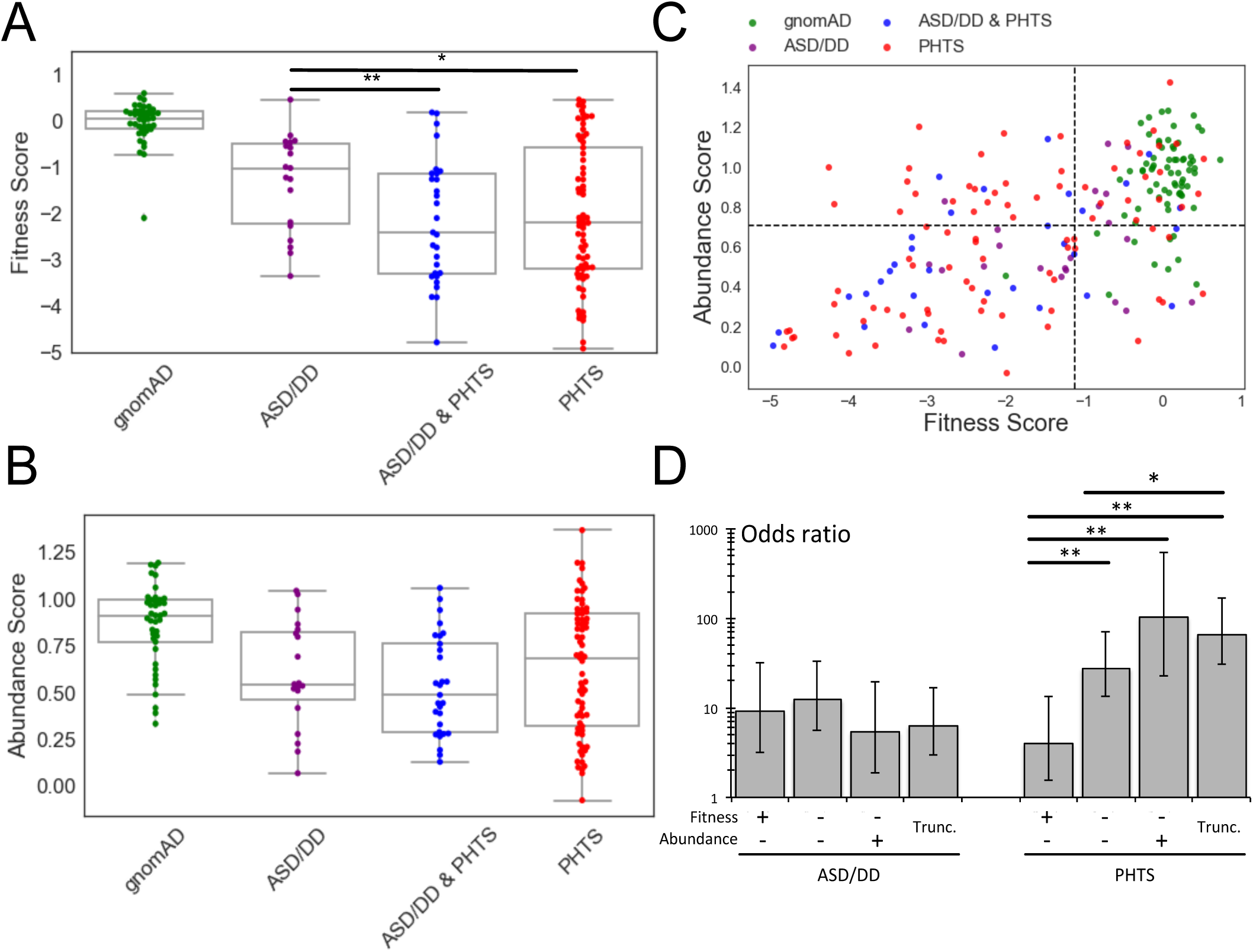
Association of molecular phenotypes with specific clinical outcomes. (A) Box plots showing fitness scores of PTEN variants occurring in individuals in gnomAD or different clinical categories. (B) Box plots showing abundance scores of PTEN variants occurring in individuals in gnomAD or different clinical categories. (C) PTEN variants plotted according to fitness and abundance scores, colored by clinical group. Dashed lines indicate hypomorphic cutoffs (-1.11 for fitness, 0.71 for abundance). (D) Odds ratios for developing ASD/DD or PHTS symptoms for different variant classes. Odds ratios represent the comparison of that class with variants in the fitness positive, abundance positive quadrant (top right in C). Error bars represent 95% confidence intervals. *p<0.05, **p<0.01.

Next, we tested whether the severity of variant molecular phenotype, as assessed by fitness or abundance score, affected the odds of developing ASD/DD or PHTS symptoms (regardless of presence or absence of the other qualifying symptoms). We included all members of the CC cohort as well as gnomAD individuals (Materials & Methods). Using a logistic regression model, we calculated odds ratios (OR) for ASD/DD and PHTS as a function of fitness or abundance scores. Wildtype-like variants were used as the reference group. For both molecular phenotypes, more severe missense variants do not significantly increase the odds of an individual developing ASD/DD (OR ranges = 3.9-6.1 and 4.2-7.8 for fitness and abundance, respectively). The odds for true truncation variants are marginally decreased compared to the missense variant classes, though this trend is not significant (Figure S4B). In contrast, for fitness and abundance scores, the odds of an individual developing qualifying symptoms for a PHTS classification increase as mutation severity increases in a stepwise manner, with stronger differences in risk observed for the abundance score (OR ranges = 20.4-51.3 and 5.1-28.7 for fitness and abundance, respectively, Figure S4B).

We next tested whether two-dimensional molecular phenotype data would provide additional insights into the risk for developing ASD/DD or PHTS symptoms (Figure 5D). While variants from the control individuals from gnomAD clearly cluster in the fitness positive, abundance positive quadrant, the affected individuals populate the other three quadrants. Interestingly, compared to the PHTS positive categories, the ASD/DD has a larger fraction of individuals in the fitness positive, abundance positive (30% vs. 10% and 20% for ASD/DD vs ASD/DD & PHTS and PHTS, respectively), and a smaller fraction in the putatively dominant negative fitness compromised, abundance positive quadrant (5% versus 21% and 25% for ASD/DD vs ASD/DD & PHTS and PHTS, respectively, Figure S4A).

Again, using a logistic regression approach, we generated odds ratios for the combined two-dimensional molecular phenotypes. We again observed no major differences in the odds for developing ASD/DD in any missense groups or the true truncation category (OR range = 5.4-12.4). In contrast, the odds for developing PHTS are highly dependent on the variant grouping (OR range = 4.1-102.9). Missense variants that maintain lipid phosphatase activity but are low abundance show the lowest odds for an individual developing qualifying symptoms for PHTS classification (OR = 4.1, 95% CI = 1.5-10.7, Figure 5D). Missense variants that were fitness and abundance negative showed a significantly different intermediate risk (OR = 27.6, 95% CI = 13.5-56.5). Variants that have wildtype-like abundance but abrogated lipid phosphatase activity (putative dominant negative variants) have the highest odds of an individual developing qualifying symptoms for PHTS classification (OR = 102.9, 95% CI = 22.8-464.0), though not significantly different from variants in the fitness negative/abundance negative or true truncation categories (Figure 5D).

## Discussion

Despite two decades of effort, we still lack a clear understanding of how *PTEN* genotype affects specific clinical phenotypes. Recent advances in DNA synthesis and sequencing technologies allow a new experimental paradigm in which the effects of thousands of variations on protein function can be empirically measured in parallel. Two such experiments recently explored the effects of *PTEN* variation on lipid phosphatase activity (fitness score) and steady state cellular abundance (abundance score).^8,12^ Using imputation, we generated estimated functional scores for all possible *PTEN* missense variants. In order to understand how molecular phenotype data relates to clinical outcomes, we integrated these data with clinical information from the CC cohort of *PTEN* mutation-positive individuals. These analyses have validated the clinical utility of comprehensive multi-dimensional functional scores and have uncovered unexpected insights into the PTEN genotype-phenotype map.

Our analyses demonstrate that molecular phenotype scores are correlated with quantitative clinical traits. Fitness and abundance scores showed a logarithmic relationship with the most penetrant *PTEN* phenotype, macrocephaly (∼95% of PTEN patients).^23^ In previous work, we designed an algorithm to determine a patient’s *a priori* risk for having a germline *PTEN* mutation (CC score). CC score is also a surrogate measure of an adult patient’s phenotypic burden accounting for age of onset.^16^ CC scores and functional scores have a linear relationship with more severe phenotypic burden associating with worse functional scores. We then demonstrated that molecular phenotype data can be used to model and thus predict likely pathogenic variants with high accuracy, compared to completely *in silico* approaches.

While broadly predicting pathogenicity has value in a clinical setting and can help resolve *PTEN* variants of uncertain significance (VUS), we were also interested in exploring if these molecular phenotypes could provide additional insights into the diverse clinical outcomes associated with germline PTEN disruption. Our analyses showed that molecular phenotypes can define subgroups of patients with common or unique age-related cancer hazard. While putative true truncating variants, such as nonsense mutations, showed high lifetime cancer risk, highly damaging missense variants as defined by the molecular phenotypes appear to be at least as impactful. Moreover, our data from single molecular phenotypes show truncation-like and true truncations survival functions separate from wildtype-like functions over an early-onset age range. Hypomorphic functions are potentially intermediate over this early on-set range but not yet significantly different from the wildtype-like functions. Combining molecular phenotype scores provides further granularity for these cancer risks and identified variants that are lipid phosphatase active (fitness positive) but unstable (abundance negative) as a significantly different intermediate class of cancer risk variants.

A growing number of studies have provided important insight into the question of genotypes driving diverse phenotypic outcomes for carriers of germline *PTEN* variants. However, these studies have generally been limited by small sample sizes. For instance, Spinelli et al. (2015) investigated the lipid phosphatase activity and protein stability of seven ASD-associated and five PHTS-associated *PTEN* missense variants using virally transfected U87MG cells. They found ASD-associated variants retained partial phosphatase function but exhibited dramatically decreased stability, whereas PHTS-associated variants lost phosphatase function but exhibited relatively better stability.^7^ These findings form the basis for the hypothesis formulated by Leslie and Longy (2016), in which ASD/DD results from hypomorphic *PTEN* variants while traditional PHTS (i.e. hamatomatous and malignant growth) results from more damaging variants.^11^ Our previous work using fitness scores (i.e. inferred lipid phosphatase activity) and ASD/DD-or PHTS-associated variants from the literature lent support to this hypothesis.^8^

Here, using the largest set of clinically annotated variants examined to date, we strengthen these previous findings by showing that ASD/DD-associated *PTEN* variants, on average, retain hypomorphic lipid phosphatase activity, while those associated with either ASD/DD and PHTS or PHTS alone are more damaging. Moreover, the fraction of missense variants and the distribution of variants according to fitness and abundance scores are more similar between the two PHTS associated groups, suggesting that they are in fact molecularly similar. We made the surprising discovery, however, that risk for developing ASD/DD is not dramatically altered across different variant loss-of-function categories, while the risk for PHTS can increase by an order of magnitude. Thus, it appears that while all individuals with pathogenic *PTEN* variants are at substantial risk for developing ASD/DD, the risk, and thus the subsequent penetrance, of PHTS symptoms (i.e. hamartomatous and malignant growth) is significantly greater for true truncations and truncation-like missense variants. These differential risk profiles would then explain the lower fraction of true truncations in cohorts recruited primarily based on an ASD/DD diagnosis.^11,19^

The biologic basis of these differential risk profiles remains unclear. Retaining any lipid phosphatase activity of the variant allele, coupled with the second functional *PTEN* allele, may be sufficient to prevent the formation of hamartomas in some cases. There are numerous PTEN functions that are not described by molecular phenotypes included in this study. Lipid phosphatase independent functions may also modulate risk. For example, recent studies have shown a potential relationship between PTEN localization and clinical outcomes, where PTEN variants showing nuclear depletion associated with ASD/DD.^24–27^ Ideally, a comprehensive analysis would include the effect of variation on PTEN’s protein phosphatase activity, subcellular localization, nuclear function, and protein-protein interaction. New high-throughput assays may make such datasets available in the near future.

Given that the majority of ASD/DD diagnoses are from children or young adults, an important open question is what will their lifetime risk for neoplasia truly be? Longitudinal tracking to definitively assess neoplasia/cancer risk in this cohort will improve the allocation of clinical resources and guide the precision delivery of care. Our current data suggest that certain subsets of individuals with *PTEN* associated-ASD/DD are likely to have higher cancer risk than other subsets. Longitudinal follow-up with these individuals or new prospective recruitment efforts will be needed to answer this question definitively.

## Data Availability

Data is available within the manuscript.

## Supplemental Data

Supplemental Data includes four figures and six tables.

## Declaration of Interests

The authors declare no competing interests.

### Acknowledgments

We thank A. C. Adey, D. M. Fowler, K. A. Matreyek, J. Zonana, G. Mandel, P. J. Stork, K. M. Wright, I. N. Smith and M. Seyfi for helpful discussions. We thank Martha Atherton and the Atherton Foundation for their support of the NARSAD awards. This work was supported, in part, by a NARSAD Young Investigator Grant from the Brain and Behavior Research Foundation through the NARSAD-Atherton Foundation Young Investigator Award (22935 to B. J. O.), a Sloan Research Fellowship in Neurosciences (Alfred P. Sloan Foundation, FG-2015-65608 to B. J. O.), the Ambrose Monell Foundation (to C.E.), the Zacconi Program of PTEN Research Excellence (to C.E.), and internal funds (C.E., B. J. O.). T. L. M received support from the Eunice Kennedy Shriver National Institute of Child Health & Human Development (F31HD095571). T. L. M. is an ARCS scholar (Achievement Rewards for College Scientists Foundation, Inc., Oregon Chapter) and B. J. O. is a Klingenstein-Simons Fellow (Esther A. & Joseph Klingenstein Fund, Simons Foundation). C. E. is the Sondra J. and Stephen R. Hardis Endowed Chair of Cancer Genomic Medicine at the Cleveland Clinic, and is an ACS Clinical Research Professor.

## Web Resources

CADD, https://cadd.gs.washington.edu/

ClinVar database, https://www.clinicalgenome.org/data-sharing/clinvar/

Consurf, http://consurfdb.tau.ac.il/

gnomAD database, https://gnomad.broadinstitute.org/downloads/

OMIM, http://www.omim.org/

PolyPhen-2, http://genetics.bwh.harvard.edu/pph2/

Provean, http://provean.jcvi.org/

STRIDE, http://webclu.bio.wzw.tum.de/stride/

## References

1. Yehia, L., and Eng, C. (2018). 65 YEARS OF THE DOUBLE HELIX: One gene, many endocrine and metabolic syndromes: PTEN-opathies and precision medicine. Endocrine-Related Cancer 25, T121–T140.

2. Yehia, L., Ngeow, J., and Eng, C. (2019). PTEN-opathies: from biological insights to evidence-based precision medicine. J Clin Invest 129, 452–464.

3. Liaw, D., Marsh, D.J., Li, J., Dahia, P.L., Wang, S.I., Zheng, Z., Bose, S., Call, K.M., Tsou, H.C., Peacocke, M., et al. (1997). Germline mutations of the PTEN gene in Cowden disease, an inherited breast and thyroid cancer syndrome. Nat. Genet. 16, 64–67.

4. Marsh, D.J., Dahia, P.L.M., Zheng, Z., Liaw, D., Parsons, R., Gorlin, R.J., and Eng, C. (1997). Germline mutations in PTEN are present in Bannayan-Zonana syndrome. Nature Genetics 16, 333.

5. Butler, M., Dasouki, M., Zhou, X., Talebizadeh, Z., Brown, M., Takahashi, T., Miles, J., Wang, C., Stratton, R., Pilarski, R., et al. (2005). Subset of individuals with autism spectrum disorders and extreme macrocephaly associated with germline PTEN tumour suppressor gene mutations. J Med Genet 42, 318–321.

6. Smith, I.N., Thacker, S., Jaini, R., and Eng, C. (2019). Dynamics and structural stability effects of germline PTEN mutations associated with cancer versus autism phenotypes. J. Biomol. Struct. Dyn. 37, 1766–1782.

7. Spinelli, L., Black, F.M., Berg, J.N., Eickholt, B.J., and Leslie, N.R. (2015). Functionally distinct groups of inherited PTEN mutations in autism and tumour syndromes. J Med Genet 52, 128–134.

8. Mighell, T.L., Evans-Dutson, S., and O’Roak, B.J. (2018). A Saturation Mutagenesis Approach to Understanding PTEN Lipid Phosphatase Activity and Genotype-Phenotype Relationships. Am J Hum Genet 102, 943–955.

9. Orloff, M.S., and Eng, C. (2008). Genetic and phenotypic heterogeneity in the PTEN hamartoma tumour syndrome. Oncogene 27, 5387–5397.

10. Mester, J., and Eng, C. (2013). When Overgrowth Bumps Into Cancer: The PTEN-Opathies. American Journal of Medical Genetics Part C: Seminars in Medical Genetics 163, 114–121.

11. Leslie, N.R., and Longy, M. (2016). Inherited PTEN mutations and the prediction of phenotype. Seminars in Cell & Developmental Biology 52, 30–38.

12. Matreyek, K.A., Starita, L.M., Stephany, J.J., Martin, B., Chiasson, M.A., Gray, V.E., Kircher, M., Khechaduri, A., Dines, J.N., Hause, R.J., et al. (2018). Multiplex assessment of protein variant abundance by massively parallel sequencing. Nature Genetics 50, 874.

13. Landrum, M.J., Lee, J.M., Riley, G.R., Jang, W., Rubinstein, W.S., Church, D.M., and Maglott, D.R. (2014). ClinVar: public archive of relationships among sequence variation and human phenotype. Nucleic Acids Res 42, D980–D985.

14. Rodríguez-Escudero, I., Roelants, F.M., Thorner, J., Nombela, C., Molina, M., and Cid, V.J. (2005). Reconstitution of the mammalian PI3K/PTEN/Akt pathway in yeast. Biochem. J. 390, 613–623.

15. Karczewski, K.J., Francioli, L.C., Tiao, G., Cummings, B.B., Alföldi, J., Wang, Q., Collins, R.L., Laricchia, K.M., Ganna, A., Birnbaum, D.P., et al. (2019). Variation across 141,456 human exomes and genomes reveals the spectrum of loss-of-function intolerance across human protein-coding genes. BioRxiv 531210.

16. Tan, M.-H., Mester, J., Peterson, C., Yang, Y., Chen, J.-L., Rybicki, L.A., Milas, K., Pederson, H., Remzi, B., Orloff, M.S., et al. (2011). A Clinical Scoring System for Selection of Patients for PTEN Mutation Testing Is Proposed on the Basis of a Prospective Study of 3042 Probands. Am J Hum Genet 88, 42–56.

17. Roche, A.F., Mukherjee, D., Guo, S.M., and Moore, W.M. (1987). Head circumference reference data: birth to 18 years. Pediatrics 79, 706–712.

18. Marsh, D.J., Kum, J.B., Lunetta, K.L., Bennett, M.J., Gorlin, R.J., Ahmed, S.F., Bodurtha, J., Crowe, C., Curtis, M.A., Dasouki, M., et al. (1999). PTEN mutation spectrum and genotype-phenotype correlations in Bannayan-Riley-Ruvalcaba syndrome suggest a single entity with Cowden syndrome. Hum. Mol. Genet. 8, 1461–1472.

19. Tan, M.-H., Mester, J.L., Ngeow, J., Rybicki, L.A., Orloff, M.S., and Eng, C. (2012). Lifetime Cancer Risks in Individuals with Germline PTEN Mutations. Clin Cancer Res 18, 400–407.

20. Saunders C.T. and Baker D (2002). Evaluation of structural and evolutionary contributions to deleterious mutation prediction. J Mol Biol. 322(4), 891–901.

21. Rentzsch, P., Witten, D., Cooper, G.M., Shendure, J., and Kircher, M. (2019). CADD: predicting the deleteriousness of variants throughout the human genome. Nucleic Acids Res 47, D886–D894.

22. Kircher, M., Witten, D.M., Jain, P., O’Roak, B.J., Cooper, G.M., and Shendure, J. (2014). A general framework for estimating the relative pathogenicity of human genetic variants. Nature Genetics 46, 310–315.

23. Mester, J.L., Tilot, A.K., Rybicki, L.A., Frazier, T.W., and Eng, C. (2011). Analysis of prevalence and degree of macrocephaly in patients with germline PTEN mutations and of brain weight in Pten knock-in murine model. Eur. J. Hum. Genet. 19, 763–768.

24. Frazier, T.W., Embacher, R., Tilot, A.K., Koenig, K., Mester, J., and Eng, C. (2015). Molecular and Phenotypic Abnormalities in Individuals with Germline Heterozygous PTEN Mutations and Autism. Mol Psychiatry 20, 1132–1138.

25. Tilot, A.K., Bebek, G., Niazi, F., Altemus, J., Romigh, T., Frazier, T.W., and Eng, C. (2016). Neural transcriptome of constitutional Pten dysfunction in mice and its relevance to human idiopathic Autism Spectrum Disorder. Mol Psychiatry 21, 118–125.

26. Tilot, A.K., Gaugler, M.K., Yu, Q., Romigh, T., Yu, W., Miller, R.H., Frazier, T.W., and Eng, C. (2014). Germline disruption of Pten localization causes enhanced sex-dependent social motivation and increased glial production. Hum Mol Genet 23, 3212–3227.

27. Fricano-Kugler, C.J., Getz, S.A., Williams, M.R., Zurawel, A.A., DeSpenza, T., Frazel, P.W., Li, M., O’Malley, A.J., Moen, E.L., and Luikart, B.W. (2018). Nuclear Excluded Autism-Associated Phosphatase and Tensin Homolog Mutations Dysregulate Neuronal Growth. Biol. Psychiatry 84, 265–277.

